# Deep learning models for COVID-19 chest x-ray classification: Preventing shortcut learning using feature disentanglement

**DOI:** 10.1101/2021.02.11.20196766

**Authors:** Caleb Robinson, Anusua Trivedi, Marian Blazes, Anthony Ortiz, Jocelyn Desbiens, Sunil Gupta, Rahul Dodhia, Pavan K. Bhatraju, W. Conrad Liles, Aaron Lee, Jayashree Kalpathy-Cramer, Juan M. Lavista Ferres

## Abstract

In response to the COVID-19 global pandemic, recent research has proposed creating deep learning based models that use chest radiographs (CXRs) in a variety of clinical tasks to help manage the crisis. However, the size of existing datasets of CXRs from COVID-19+ patients are relatively small, and researchers often pool CXR data from multiple sources, for example, using different x-ray machines in various patient populations under different clinical scenarios. Deep learning models trained on such datasets have been shown to overfit to erroneous features instead of learning pulmonary characteristics – a phenomenon known as shortcut learning. We propose adding feature disentanglement to the training process, forcing the models to identify pulmonary features from the images while penalizing them for learning features that can discriminate between the original datasets that the images come from. We find that models trained in this way indeed have better generalization performance on unseen data; in the best case we found that it improved AUC by 0.13 on held out data. We further find that this outperforms masking out non-lung parts of the CXRs and performing histogram equalization, both of which are recently proposed methods for removing biases in CXR datasets.

## 1 Introduction

The Coronavirus Disease (COVID)-19 pandemic has exposed many vulnerabilities in public health systems around the world. Artificial intelligence (AI) is poised to address some of these challenges with potential applications including early disease detection, clinical management tools, disease modeling, and vaccine research^1^. Screening patients for COVID-19 based on chest radiograph (CXR) imaging is one potential AI application, and deep learning models have been developed to distinguish COVID-19 pneumonia from normal findings and even rule out other types of pneumonia with high accuracy ^2–4^. Due to the increasing practice of reporting results prior to peer review in the setting of a novel pandemic, many additional models have been reported as preprint papers^5–11.^

The main challenge with training these deep learning models has been the shortage of COVID-19 CXR imaging data. Several public datasets are available, but most are small (100-200 patients). A recent review assessed many AI diagnostic and prognostic models for COVID-19 using the prediction model risk of bias assessment tool (PROBAST) and found that all but one of the 51 studies were at high risk for analysis bias, mainly overfitting due to small sample size^12^. Larger databases have typically pooled images from different patient populations at multiple research sites and hospitals, or even from different countries^7,13,14.^ These unbalanced datasets often contain images with source-specific identifiers (anteroposterior versus posteroanterior positioning, imaging device type, image size, etc) that the models can misidentify as relevant features.

The most common approach for developing COVID-19 CXR diagnostic models is to use transfer and representational learning techniques to adapt classification models that were trained on labeled CXR datasets (to classify pre-COVID-19 pulmonary findings) or on unrelated image datasets to the new task^2^,^4,15^. However, due to the problems mentioned above – small datasets or larger datasets with a latent imbalance – leads to learned models that suffer from a phenomenon summarized as “shortcut learning”^16^. Here, the models will identify dataset-specific features rather than pulmonary specific features in order to distinguish between classes, and the reported performance of these models may not be generalizable to other patient populations. Indeed, recent research proves exactly this^17^ and further shows that some shortcuts can persist in external datasets. The problem now is in how to train highly-parameterized deep learning models on publicly available data, without learning shortcuts that are present in such data. A recent study proposed two preprocessing techniques to solve this problem: histogram equalization, to correct for issues such as contrast differences between images, and lung masking to remove potential source-specific information located outside of the lung area^18^. See Supplemental Information 1 for an expanded literature review of work that uses CXRs for COVID-19, domain adaptation and disentangled representation learning.

In this paper, we sought to evaluate the use of feature disentanglement^19^, a multi-task training approach for deep neural networks, to prevent shortcut learning in the context of automated classification of COVID-19 CXR images. Models trained with this method are forced to learn features that can identify COVID-19+ CXRs and are penalized for learning features that can identify what sub-population the CXR is from. We further sought to compare feature disentanglement to the previously proposed histogram equalization and and lung masking methods. Finally, we sought to test all three methods on the ultimate goal of improving model generalization performance on unseen data. We have released source code to reproduce the models and results found in this paper: https://github.com/microsoft/xray-feature-disentanglement.

## 2 Methods

### 2.1 Datasets

We used two CXR datasets: the open-source COVIDX dataset^20^ and a dataset received from the China Consortium of Chest CT CC-CCII^21^. These datasets share the same three *class* labels for CXR images: “normal” (Normal) – collected from patients without pneumonia, “common pneumonia” (Pneumonia) CXRs – collected from patients with pneumonias other than from COVID-19, and “novel COVID-19 pneumonia” (COVID-19+). We treated the source dataset of each image as its *domain* label. See Table 1 for a breakdown of the number of *class* labels of each type per dataset. Below we describe each dataset in more detail.

**Table 1.** Dataset overview. Counts of disease label type per dataset.

|  | Normal | Pneumonia | COVID-19+ |
| --- | --- | --- | --- |
| **COVIDx (totals)** | 8,851 | 6,040 | 421 |
| COVID-CHESTXRAY | – | 28 | 289 |
| FIGURE 1 | – | – | 35 |
| ACTUALMED | – | – | 51 |
| SIRM | – | – | 46 |
| RSNA | 8,851 | 6,012 | – |
| **CC-CCII** | 11,604 | 18,236 | 1,690 |

The COVIDx dataset is a conglomeration of samples from five other datasets: the COVID-CHESTXRAY dataset (also referred to as the COHEN dataset in related work)^22^, the FIGURE 1 COVID-19 CXR dataset^23^, the ACTUALMED COVID-19 CXR dataset^24^, the Kaggle RSNA Pneumonia Detection Challenge dataset^25^, and the SIRM samples from the Kaggle COVID-19 radiography database^26^. The samples are divided among three classes: “normal” control images, non COVID-19 pneumonia images, and COVID-19+ images. We used the scripts provided on the COVIDx GitHub repository to create the dataset. We then merge the training and test sets and filter out all but the first sample for each patient. The label counts in Table 1 for this dataset therefore equal the number of patients.

**Figure 1.**
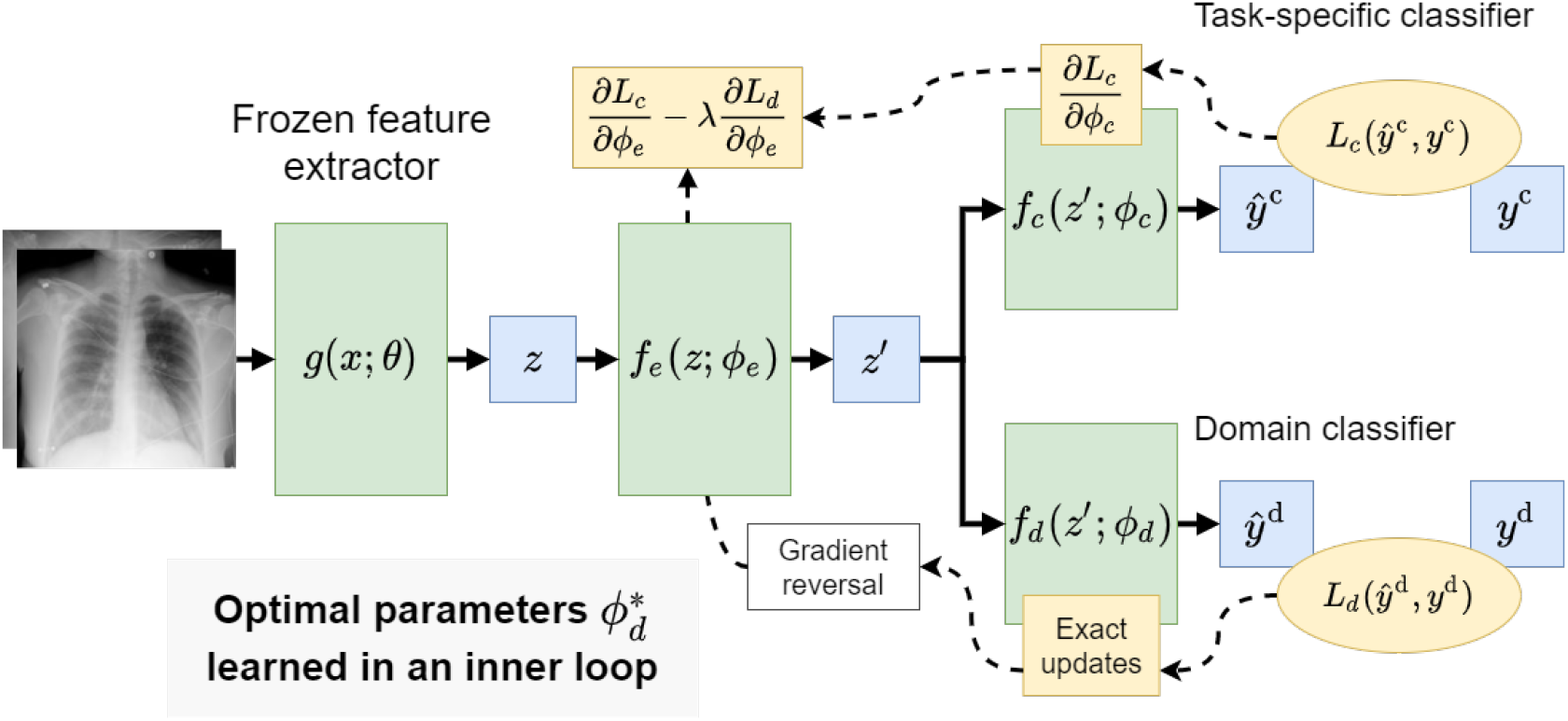
Overview of the feature disentanglement modeling approach. We propose to learn a model that simultaneously predicts the class label and domain label for a given CXR image. The parameters of the model are updated to extract representations that contain information about the class label but not about domain label.

The CC-CCII dataset is a large CT dataset encompassing CT images from retrospective cohorts from the China Consortium of Chest CT Image Investigation (CFC-CCII)^21^. This datasets has samples from Sun Yat-sen Memorial Hospital and Third Affiliated Hospital of Sun Yat-sen University, The First Affiliated Hospital of Anhui Medical University, West China Hospital, Guangzhou Medical University First Affiliated Hospital, Nanjing Renmin Hospital, Yichang Central People’s Hospital, and Renmin Hospital of Wuhan University. The dataset we use in this paper consists of CXRs taken from a subset of the patients included in the CT dataset. These images have been labeled with the same classes as in COVIDx.

#### Ethics Statement

This study was conducted in accordance with the Declaration of Helsinki. The COVIDx dataset is publicly available. The CC-CCII dataset was de-identified and anonymized and this study was retrospectively approved by the institutional review board of Sun Yat-sen Memorial Hospital.

### 2.2 Model training

Formally, we trained a model, *f* (*g*(*x*; *θ*); *ϕ*), where *x* is a CXR. This model is decomposed into a feature-extractor, *g*(*x*; *θ*) = *z*, parameterized by *θ*, and classifier, *f* (*z*; *ϕ*), parameterized by *ϕ*, where the feature-extractor is responsible for creating an embedding, *z*, for a given CXR, and the classifier is responsible for predicting the label from a given embedding. This representation is helpful in transfer learning settings – frozen feature-extractor models (i.e. models with fixed *θ*) that have been pre-trained on large CXR datasets can be used to reduce overfitting when relatively few labeled samples are available.

We fit *f* (*z, ϕ*) over a dataset, 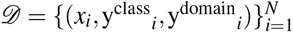, where each image contains a *class* label and a *domain* label. The *class* labels are clinically relevant; for example, they describe whether a CXR is from a normal (Normal) patient, a patient with common pneumonia (Pneumonia), or from a patient that may exhibit novel COVID-19 pneumonia (COVID-19+). The *domain* labels are *not* clinically relevant; they encode information about how the sample was collected, such as the clinical site and patient cohorts. In our experiments, the *domain* labels encode which dataset a sample originally came from. An ideal classifier would not be sensitive to how a sample was collected but would rely on pulmonary features present in the different types of *class* labels. However, for the reasons we outline in the Introduction, classifiers can overfit to spurious signals in CXRs leading to poor generalization to new imagery.

We fit *f* (*z*; *ϕ*) in two optimization settings: a straightforward baseline setting and a feature disentanglement setting in which we force the classifier to learn data representations that are not useful in predicting the *domain* labels, while remaining useful for predicting the *class* labels:

#### Baseline

In this setting, we learned *ϕ*^*∗*^ by minimizing a negative log-likelihood loss, *L*(ŷ^class^, y^class^), between the class predictions, ŷ^class^, and the *class* labels, y^class^, over *𝒟* in a standard setup:

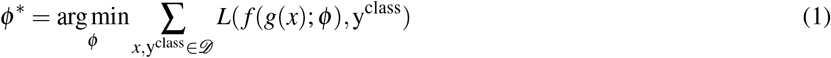

Our hypothesis was that, in this setting, even simple classifiers will overfit to spurious features and exhibit poor generalization performance.

#### Feature disentanglement

In this setting we assumed that we can further decompose *f* into a feature extractor, 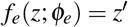, and two classification heads, 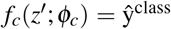 and 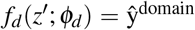 – see Figure 1 for an overview of this setup. We then follow the methodology proposed in^19^ for training *f* in a multi-task setting to transform the initial embedding *z* into a compressed form *z*^*′*^ that is useful for predicting the class label and not useful for predicting the domain label for a given CXR. Formally, we defined the empirical error with a *class* loss as above, and an additional *domain* loss:

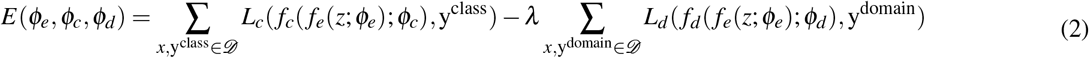

Note that the error is defined as the difference between the summed class loss and summed domain loss. Larger loss values in the second term will contribute to the overall minimization of the total error which influence the model to learn a compressed representation that is *not* predictive of the domain labels. *λ* is a hyperparameter that controls the relative influence of the domain loss on the error. We optimized for parameters, *ϕ*, according to:

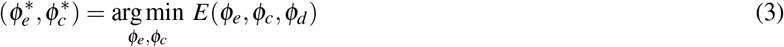

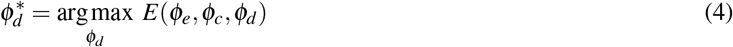

As described in^19^, this can be solved by iterating using gradient-based optimization with an additional “gradient reversal layer” inserted between *f*_*e*_ and *f*_*d*_ in the structure of the model (see Figure 1). We considered a modification wherein after each training epoch we optimize for 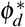, over the whole training set, for the current, fixed values of *ϕ*_*e*_. Put simply, we trained 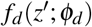 to convergence at the end of each epoch. As a consequence, the updates to *ϕ*_*e*_ in the next epoch of training are done with respect to the domain loss computed at a local minimum for the domain classifier. We found empirically that this helped reduce variance in results between training runs. This modification is cheap when the parameters of the original feature extractor remain frozen and the intermediate representations, *z*, are cached – which was always the case in our experiments.

### 2.3 Lung masking and histogram equalization preprocessing

Motivated by the same points that we are,^18^ proposed two preprocessing methods for eliminating signal in CXR imagery that a classifier might overfit to: lung masking and histogram equalization. Lung masking uses a model to remove the non-lung parts of CXR images with the assumption that features from the imagery surrounding the lungs should not be used in predictive models of lung diseases. Histogram equalization is a common technique in computer vision used to improve the contrast of grayscale images. If histogram equalization is applied to each image in a dataset of CXR image, then a classification model will not be able to exploit relative differences in the contrast of images in its decisions.

These methods can be used in combination with our proposed method for feature disentanglement. In our experiments we used the recent SOTA lung VAE model^27^ to create lung masks and implementation of histogram equalization from OpenCV^28^.

### 2.4 Experimental setup

Our method depends on a feature extractor model, *g*(*x*; *θ*), to create an initial set of embeddings. We considered three different pre-trained CNNs for this role:

#### Torchxrayvision pretrained DenseNet121

We used the DenseNet121 model^29^ weights released in the torchxrayvision package^30, 31.^ This model has been trained on CXR imagery, collected before the COVID-19 pandemic, with multi-task pulmonary disease labels from different datasets. We extracted a 1,024 dimensional feature vector for a given CXR by applying global average pooling after the last convolutional layer in the DenseNet.

#### ImageNet pretrained DenseNet121

We also used the DenseNet121 model^29^ weights from the PyTorch torchvision library^32^. This model has been trained on ImageNet, and the *only* difference in implementation from the torchxrayvision version is that the first convolutional layer operates over 3 channel images instead of 1 channel images. We duplicated input values across the 3 input channels in order to run this model on CXR imagery. We extract features from this DenseNet in the same way as above.

#### COVID-Net

We used the COVID-Net model and the “COVIDNet-CXR Large” pretrained weights from the same repository that proposed the COVIDx dataset^20^. We extract a 2,048 dimensional feature vector for an input CXR by applying global average pooling after the last convolutional layer in the COVID-Net architecture. This is in contrast with the methodology from^20^, who flatten the representation after the last convolutional layer to get a 460,800 dimensional representation.

In all experiments we reported average and standard deviation results from 5*×* 10-fold cross validation. In Section 3.1 we stratified by the domain label in order to ensure that there are samples from each dataset in training and testing splits. In Section 3.2 where we report results on the out-of-sample CC-CCII data, we used the out-of-sample data as an additional test set in the cross-validation folds used to test in-sample performance. Specifically, we split the training set into 10-folds, trained a model on data from 9 folds, then tested on the held-out fold, as well as the entire out-of-sample dataset.

Our classification problems are all multi-class; we reported an unweighted average of the area under the ROC curve (AUC) calculated individually for each class in a one-versus-rest manner and average per class accuracy (ACC). Both methods are not sensitive to the class imbalance which we observe in both our *class* and *domain* tasks.

When using feature disentanglement, we set *f*_*e*_(*z*) as a fully connected model with the following structure: Dense(256) *→* BatchNorm*→* Dense(64) *→* BatchNorm, where both dense layers are followed by a ReLU nonlinearity. We set both *f*_*c*_(*z*^*′*^) and *f*_*d*_(*z*^*′*^) as logistic regression layers. We trained with the AdamW optimizer using AMSGrad and an initial learning rate of 0.001. We divided the learning rate by a factor of 10 if the validation *class* loss has stagnated for over 10 epochs and stopped training either the third time this happens, or after 200 total epochs.

We also decayed *λ* throughout training; we set *λ*_0_ = 10 and use the following update rule evaluated each epoch (where *t* is the epoch) to calculate *λ* :

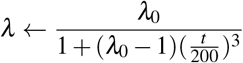

This moves *λ* from *λ*_0_ to 1 over the course of the maximum 200 epochs of training. We did not thoroughly test this schedule against other choices. We simply aimed for the domain loss to guide training in the initial epochs of training, and for the class loss to converge to a local minimum in the later epochs of training.

Finally, all of our experiments are fully reproducible by setting the seed of the random number generators in all component software packages. We varied this seed over the 5x restarts in our experiments.

## 3. Experiments and Results

### 3.1 Identifying domain labels from pre-trained model representations

In Table 2 we show that all of our existing models extract representations that contain enough information, even when run on masked/equalized imagery, for discriminating between domain labels throughout the COVIDx dataset, and domain labels throughout *only* COVID-19+ samples. For example, we find that a DenseNet121 pre-trained on ImageNet extracts features from unmasked CXR images that are sufficient to train a logistic regression model that can identify the source dataset among samples from the COVIDx dataset with a held-out average AUC of 0.95*±* 0.02. The same embeddings can be used to train a logistic regression model that can identify the source dataset given COVID-19+ sample, among the constituent datasets of COVIDx and CC-CCII with a held out AUC of 0.97 *±*0.01. While the preprocessing steps help to reduce this performance, the linear models still perform much better than random guessing, suggesting that overfitting to domain signals in the embedded representations is trivial.

**Table 2.** Results showing how well classifiers can identify which sub-dataset a CXR is from within the COVIDx dataset and how well classifiers can identify which dataset a “COVID-19+” CXR is from across both the COVIDx and CC-CCII datasets. We report AUC values as averages of the one-vs-all binary AUCs between all classes, and accuracy (ACC) as the average accuracy over all classes. We observe that the representations generated by the classifiers, even from masked/equalized inputs, contain enough information to accurately identify the sources of the imagery in both cases.

| Unmasked Images | Embedding size | COVIDx datasets |  | All COVID-19+ samples |  |
| --- | --- | --- | --- | --- | --- |
|  |  | AUC | ACC | AUC | ACC |
| Pixel intensity histogram | 256 | $0.75 \pm 0.03$ | $0.36 \pm 0.04$ | $0.83 \pm 0.03$ | $0.47 \pm 0.07$ |
| Torchxrayvision embedding | 1024 | $0.93 \pm 0.03$ | $0.54 \pm 0.09$ | $0.93 \pm 0.03$ | $0.59 \pm 0.07$ |
| ImageNet embedding | 1024 | $0.95 \pm 0.02$ | $0.53 \pm 0.07$ | $0.97 \pm 0.01$ | $0.66 \pm 0.09$ |
| COVID-Net embedding | 2048 | $0.89 \pm 0.05$ | $0.39 \pm 0.05$ | $0.91 \pm 0.03$ | $0.45 \pm 0.04$ |
| Masked/equalized Images | Embedding size | AUC | ACC | AUC | ACC |
| Pixel intensity histogram | 256 | $0.57 \pm 0.06$ | $0.27 \pm 0.06$ | $0.75 \pm 0.02$ | $0.25 \pm 0.04$ |
| Torchxrayvision embedding | 1024 | $0.76 \pm 0.05$ | $0.35 \pm 0.07$ | $0.88 \pm 0.02$ | $0.42 \pm 0.08$ |
| ImageNet embedding | 1024 | $0.84 \pm 0.02$ | $0.34 \pm 0.04$ | $0.91 \pm 0.02$ | $0.50 \pm 0.10$ |
| COVID-Net embedding | 2048 | $0.71 \pm 0.04$ | $0.26 \pm 0.04$ | $0.85 \pm 0.04$ | $0.35 \pm 0.05$ |

### 3.2 Class and domain performance

In Table 3 we show the within-dataset and generalization performance of models, *f* (*z*; *ϕ*), trained on top of the different feature extractors. Similar to Table 2, the models that are trained in a standard way have high performance on the domain task, and correspondingly high performance on the actual task labels. At the same time, the poor performance of these models on the out-of-sample dataset, CC-CCII, shows that they are overfitting to the training dataset. For example, a model trained on top of torchxrayvision unmasked image embeddings performs similar to random guessing – random guessing performance is 0.33 average accuracy, while this model gets 0.34 average accuracy. Lung masking and histogram equalization improves generalization performance in all cases, however we hypothesize that this may throw out signals that are relevant to the task.

**Table 3.** Results showing within-dataset class performance, within-dataset domain performance, and out of sample (OOS) class performance from training models with the COVIDx dataset. The class performance shows how well classifiers are able to distinguish between “Normal”, “Pneumonia”, and “COVID-19+” disease labels, while the task performance shows how well classifiers are able to distinguish which constituent dataset an image belongs to. We report AUC values as averages of the one-vs-all binary AUCs between all classes, and accuracy (ACC) as the average accuracy over all classes. In all cases class performance (both within-dataset and OOS) is reported from the classifier trained on samples within-dataset, while domain performance is reported from an additional classifier trained to predict domain labels on top of the learned representations, *z*^*′*^, as a measure of how much domain information the representation contains. We observe that using feature disentanglement decreases within-dataset domain performance as expected, and increases OOS class performance – i.e. improves generalization performance.

|  | COVIDx |  | CC-CCII |  |
| --- | --- | --- | --- | --- |
|  | Task AUC | Domain AUC | Task AUC | Task ACC |
| **Torchxrayvision Embeddings** |  |  |  |  |
| Unmasked | $0.97 \pm 0.01$ | $0.94 \pm 0.02$ | $0.55 \pm 0.03$ | $0.34 \pm 0.04$ |
| Masked/equalized | $0.92 \pm 0.01$ | $0.85 \pm 0.03$ | $0.65 \pm 0.02$ | $0.42 \pm 0.02$ |
| Unmasked + Disentanglement | $0.90 \pm 0.03$ | $0.56 \pm 0.07$ | $0.68 \pm 0.04$ | $0.49 \pm 0.02$ |
| Masked/equalized + Disentanglement | $0.87 \pm 0.02$ | $0.53 \pm 0.06$ | $0.71 \pm 0.03$ | $0.47 \pm 0.02$ |
| **ImageNet Embeddings** |  |  |  |  |
| Unmasked | $0.96 \pm 0.01$ | $0.97 \pm 0.02$ | $0.64 \pm 0.02$ | $0.37 \pm 0.01$ |
| Masked/equalized | $0.94 \pm 0.01$ | $0.88 \pm 0.03$ | $0.67 \pm 0.03$ | $0.41 \pm 0.03$ |
| Unmasked + Disentanglement | $0.85 \pm 0.03$ | $0.57 \pm 0.07$ | $0.73 \pm 0.03$ | $0.43 \pm 0.02$ |
| Masked/equalized + Disentanglement | $0.85 \pm 0.03$ | $0.57 \pm 0.04$ | $0.73 \pm 0.03$ | $0.46 \pm 0.02$ |
| **COVID-Net Embeddings** |  |  |  |  |
| Unmasked | $0.96 \pm 0.01$ | $0.93 \pm 0.02$ | $0.59 \pm 0.02$ | $0.38 \pm 0.01$ |
| Masked/equalized | $0.89 \pm 0.02$ | $0.80 \pm 0.04$ | $0.63 \pm 0.02$ | $0.44 \pm 0.02$ |
| Unmasked + Disentanglement | $0.92 \pm 0.02$ | $0.53 \pm 0.10$ | $0.67 \pm 0.03$ | $0.41 \pm 0.02$ |
| Masked/equalized + Disentanglement | $0.83 \pm 0.02$ | $0.52 \pm 0.07$ | $0.62 \pm 0.01$ | $0.45 \pm 0.00$ |

On the other hand, we observe that training *f* (*z*; *ϕ*) with feature disentanglement results in better generalization performance with both unmasked or masked/equalized images in all cases. ImageNet embeddings of masked/equalized images and feature disentanglement training results in the best generalization performance on CC-CCII. Interestingly, training with feature disentanglement on masked/equalized images is not always better than training with feature disentanglement on unmasked images. This supports the idea that lung masking and histogram equalization might throw out relevant task signals. In all cases, training with feature disentanglement dramatically reduces the domain signal in learned representations. For example, the domain performance of the model tuned with torchxrayvision embeddings of unmasked images is 0.94 AUC, while using feature disentanglement reduces this to 0.56 AUC. The increase in performance from training with feature disentanglement is variable across the pre-trained model used to generate the initial representation. We observe the smallest difference in generalization performance with the COVID-Net pre-trained model which was initially trained on unmasked images from the COVIDx dataset – a smaller dataset than either ImageNet or the CXR dataset used by torchxrayvision – and may not function as an effective feature extractor.

Further experiments are needed to determine if unfreezing the parameters of the feature extractor model during training with feature disentanglement is beneficial. We specifically avoid this as we have already found that it is possible to overfit to the training set with a small fully connected model on top of pre-trained embeddings, as well as learn representations that are not predictive of the within-dataset domain labels. Performance gains of fine-tuning through the feature extractor would only be potentially visible through increases in generalization performance, i.e. on the CC-CCII dataset. By correlating such design decisions with CC-CCII performance. We would risk manually overfitting to CC-CCII, and want to avoid doing so in this work.

### 3.3 Feature visualization

In Figure 2 we show the UMAP embeddings of the learned feature representations from the models trained on the torchxrayvision embeddings for images from the COVIDx dataset. This shows how the representations learned without feature disentanglement clearly separate the images from the RSNA dataset from the images from the other datasets, despite not being trained to do this. This is because the RSNA dataset contains most of the samples with “normal” and “pneumonia” disease labels, versus the other datasets, which contain “COVID-19+” samples. Therefore, when a model is trained to separate these classes, it finds features that are correlated with which component dataset an image is from. Such biases can be trivial, such as a different in contrast between the dark and light portions of the image, the position/size of the lungs, markings on the CXR, etc. The same distinction between datasets is not clear when feature disentanglement is used because the learned representations are forced to not be predictive of dataset. As a result, the distinction between “COVID-19+” and “pneumonia” labels becomes less clear when feature disentanglement is used, however, as we show in Table 3, this can improve the generalization ability of the model.

**Figure 2.**
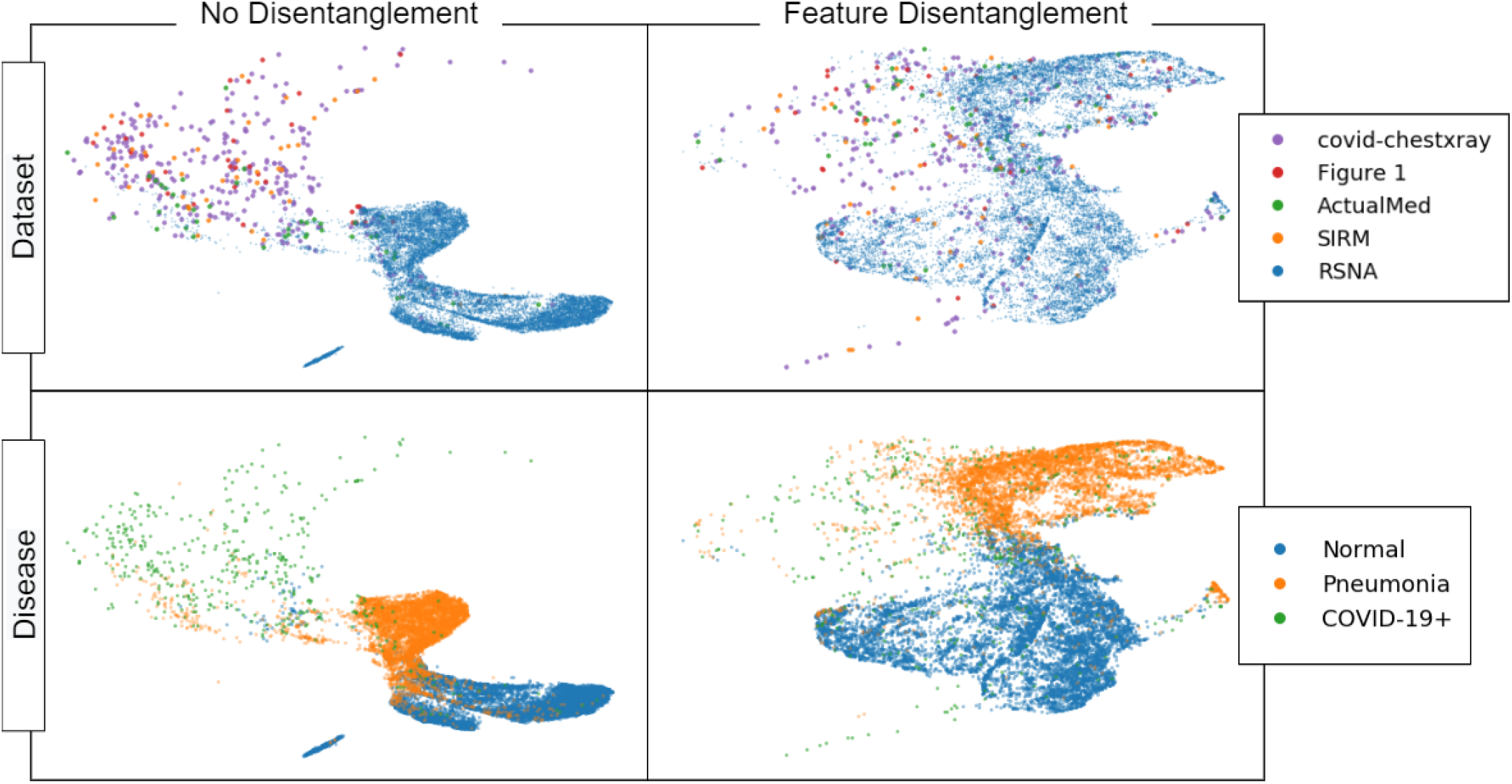
UMAP projections of features learned from models trained with and without feature disentanglement on unmasked imagery. Each point represents a CXR from the COVIDx dataset. The top row colors points by their domain label – which subdataset of the COVIDx dataset they are in – while the bottom row colors points by their disease label. We observe that without feature disentanglement, the learned representations easily separate datasets – despite not being trained for this task – however, with feature disentanglement, the learned representations do not clearly separate datasets.

## 4 Discussion

Accurate automated diagnosis of COVID-19 pneumonia based on CXRs has been a research focus since the start of the pandemic, given its potential use in emergency departments, urgent care, and resource-limited settings. The development of such devices has been limited by the availability and quality of COVID-19+ datasets, which are typically aggregated from various sources to increase the number of COVID-19+ examples. In this study we show that previously proposed techniques, such as isolating the pulmonary region and harmonizing images taken from different x-ray machines, fail to prevent deep learning models from relying on features that are specific to a particular dataset or image source (e.g. scanner machine type, age of patient, external artifacts in the image). These models are likely to underperform in real-world clinical settings, when analyzing images that lack the identifying data the models have learned to rely on.

Since the COVID-19 outbreak, various researchers have developed automated COVID-19 CXR diagnosis models. Most previous studies have used transfer learning approaches and compared classification performances obtained between several popular CNN architectures, but all have relied on datasets made up of COVID-19+ CXRs sourced from around the web^12^. For COVID-19 negative cases, data are typically sampled from other open CXR datasets. However, if any bias is present within these datasets, the model could learn the underlying biases, rather than learning COVID-19 related features. Therefore, we have presented an application of feature disentanglement for predicting COVID-19 infection even when training on public datasets containing CXRs from multiple sources. The model showed superior performance when tested on a new and unfamiliar dataset, suggesting that it was relying on COVID-19 specific pulmonary findings. We applied visualization techniques to show that our model relied on imaging features that were not specific to particular groupings of CXR images in the training data, further suggesting that the model is capable of performing well when analyze CXR data from unfamiliar sources

Although we show that our approach produces a model that is more successful when challenged with real-world data, our work is still limited by the lack of public datasets available for testing, and we used a private dataset from the CC-CCII to test generalization performance. Future work should investigate the effects of fine-tuning these models with and without feature disentanglement approaches, and the effects of controlling for multiple imaging data sources in an expanded multi-task setting. Finally, when larger CXR datasets that are paired with clinical outcomes become available, further exploration of the use of feature disentanglement for extracting small sets of clinically relevant features from CXRs should be investigated. However, the diagnostic performance of any diagnostic model should be carefully evaluated in a real-world setting before clinical implementation. Misdiagnosis of COVID-19 can lead to inappropriate care, failure to treat, increased transmission, and many other serious outcomes. Clinicians should be aware of potential limitations and biases when incorporating model predictions into their clinical assessment.

Finally, our approach has potential clinical applications beyond automated diagnosis. CXR diagnostic models that rely on relevant pulmonary findings may be also useful for the development of prognostic models, by combining the CXR information with other clinical and demographic data to predict which patients are at risk for severe disease.

## Data Availability

The COVIDx dataset is publicly available and can be found at the links provided in the "Data Availability Links" section.
The CC-CCII dataset that we use is not publicly available.

https://github.com/lindawangg/COVID-Net

## Author contributions statement

C.R. certifies that all authors labeled with “+” should be considered first-authors to all academic and professional effects, and that their names can be legitimately swapped in their respective publication lists. C.R., A.T., and A.O. conducted the experiments. M.B., A.L., C.R, P.B, and J.M.L.F. analysed the results. All authors reviewed the manuscript.

