## Supplemental Information for "Deep learning models for COVID-19 chest x-ray classification: Preventing shortcut learning using feature disentanglement"

### 1 Supplementary related work

**Using CXRs for COVID-19.** Since the COVID-19 outbreak, various research has tried COVID-19 diagnosis with Convolutional Neural Networks (CNNs) on radiographic images. Many approaches to classify chest x-ray scans to discriminate COVID-19 positive cases have been shown. Focusing on a transfer learning-based approach, (1; 2) compare various classification performances obtained between several popular CNN architectures. A similar approach employed by (3) uses Resnet-based architectures with a 5-fold cross-validation strategy. (4) propose a novel CNN architecture for the COVID classification task. However, all this research relies on the open-source COVID-CHESTXRAY dataset (5), made up of COVID-19+ CXRs sourced from around the web. For COVID-19 negative cases, data are typically sampled from other open CXR datasets. However, if any bias is present within these datasets, the model could learn the underlying biases, rather than learning COVID-19 related features. For instance, a model could potentially learn to discriminate based on differences due to the scanning devices, or unique windowing parameters of each CXR, or some other acquisition settings. This can result in the classification task yielding apparently optimal classification performance. Domain adaptation techniques like feature disentanglement can be useful to address this issue.

**Domain Adaptation.** Domain adaptation (DA) transfers the knowledge learned from one or more source domains to a target domain. Discrepancy-based DA approaches (6; 7; 8; 9), adversary-based approaches (10; 11; 12), and reconstruction-based approaches (13; 14; 15; 16) are designed to handle a single-source to single-target adaptation. Originating from the theoretical analysis in (17; 18; 19), the multiple source domain adaptation (MSDA) approach assumes that training data are collected from multiple sources and has been applied to several practical applications (20; 21; 22) (17) introduces an approach with H$\Delta$H-divergence between the weighted combination of source domains and a target domain.

**Disentangled Representation Learning.** Disentangled representations learning tries to model the factors of knowledge variation. (23; 24; 12; 25) aims at learning an interpretable representation using generative adversarial networks (GANs) (26; 27) and variational autoencoders (VAEs) (28; 29). (30) proposes to disentangle the feature representation into a domain-invariant content space and a domain-specific attribute space in a fully supervised setting. (25) proposes an auxiliary classifier GAN (AC-GAN) to achieve representation disentanglement. However, all these approaches specialize in disentangling representation in a single domain. (12) introduces a unified feature disentangler for domain-invariant representation from data across multiple domains. However, they assume multiple source domain availability during training, which limits its application.
